# Comparison of cardiovascular risk in individuals with normal vs isolated elevated diastolic blood pressure

**DOI:** 10.1101/2025.09.10.25335500

**Authors:** Maryne Lepoittevin, Pierre Bauvin, Alaedine Benani, Philippe Attias, Ph.Gabriel Steg, Emmanuelle Vidal-Petiot, Sylvain Bodard

## Abstract

**Background:** In 2024, the European Society of Cardiology (ESC) hypertension guidelines introduced an “elevated blood pressure” category (120–139/70–89 mmHg), lowering the normal diastolic BP threshold from 85 to 70 mmHg. The implications of this change for risk stratification in primary prevention are uncertain.

**Methods:** We conducted a cross-sectional study of adults undergoing standardized preventive health assessments at a dedicated center in Paris, France. Office blood pressure was measured with a validated automated oscillometric device. Participants were classified using ESC/ESH□2018 and ESCL2024 definitions. We quantified shifts across BP categories and compared clinical, lifestyle, and biological profiles between individuals reclassified from ESCL2018 “Optimal” to ESCL2024 “Elevated” solely due to diastolic BP ≥L70LmmHg with systolic BP<□120□mmHg.

**Results:** Among 1,394 participants (mean age 49.9 ± 12.1 years; 33.9% women), ESC 2024 classified 10.0% as non-elevated (<120/70 mmHg), 64.2% as elevated, and 25.8% as hypertensive (≥140/90 mmHg). Overall, 328 (23.5%) moved from ESC/ESH2018 “Optimal” (BP <120/80 mmHg) to ESC 2024 “Elevated” on the basis of diastolic pressure alone. Compared with individuals classified in the 2018 optimal and 2024 non-elevated subgroup (BP <120/70 mmHg), reclassified participants (systolic BP <120 and diastolic BP 70– 79 mmHg) were modestly older (45.5 vs 42.7 years; p = 0.007) but did not differ by sex, body-mass index, smoking exposure, alcohol consumption, self-rated health, cardiovascular history, or routine biomarkers. SCORE2 did not differ between these groups (p = 0.12), but increased progressively across successively higher ESC/ESH2018 categories. In line with this gradient, ESC/ESH2018 “Optimal” versus non-optimal groups differed significantly across multiple risk markers (all p < 0.05).

**Conclusions:** In this low-risk preventive cohort, lowering the diastolic threshold to 70 mmHg reclassified nearly one quarter of adults with previously optimal BP into the elevated BP category, without identifying a clinically distinct higher-risk phenotype. Prospective studies with adjudicated outcomes are needed to determine the utility of this threshold for primary prevention.

## Introduction

Arterial hypertension remains the leading modifiable risk factor for cardiovascular morbidity and mortality worldwide, accounting for over 10 million deaths annually (1–3). In this context, clinical guidelines play a central role in defining blood pressure (BP) thresholds for diagnosis, risk stratification, and treatment decisions. In 2024, the European Society of Cardiology (ESC) introduced a new simplified blood pressure classification, including a new blood pressure category, termed “elevated blood pressure”, and defined as office systolic BP between 120 and 139 mmHg and/or diastolic BP between 70 and 89 mmHg (4). Non-elevated BP is the category below 120/70 mmHg, while the definition of hypertension is unchanged, ≥ 140/90 mmHg. This three-category classification replaces the previous gradation used in the 2018 ESC/ESH guidelines, which, below the threshold defining hypertension, distinguished between “optimal” (<120/80 mmHg), “normal” (120–129/80–84 mmHg), and “high-normal” (130–139/85–89 mmHg) BP (5). As a result, individuals with values such as 118/72 mmHg, previously classified as “optimal,” are now labeled as having elevated BP, as are all those with previously “normal” BP (5).

The downward shift in diastolic blood pressure threshold, from 85 mmHg (the upper limit of normal diastolic BP in the 2018 ESC/ESH classification) to 70 mmHg in the 2024 guidelines, represents a substantial conceptual and clinical change. While the reduction of the systolic threshold to 120 mmHg is supported by epidemiological data showing a continuous increase in cardiovascular risk from systolic BP values starting below 120 mmHg (6,7) and by interventional trials showing improved outcomes when targeting a lower SBP (systolic BP), the decision to lower the diastolic threshold is not supported by comparable evidence. In fact, several large-scale observational studies have failed to demonstrate a consistent association between DBP (diastolic BP) below 80mmHg and adverse cardiovascular outcomes (8). Some analyses even suggest a U-shaped relationship, with excessively low diastolic BP potentially associated with increased coronary risk, particularly in older individuals or those with underlying atherosclerosis (10).

A recent analysis using data from the French CONSTANCES cohort showed that only 16□% of adults aged 18–69 were categorized as having “non-elevated” BP under the 2024 ESC criteria, compared with more than 50□% with “optimal” or “normal” BP under the 2018 classification (11). This reclassification would lead to a dramatic increase in the population labeled as having abnormal BP, with the potential for treatment initiation in some individuals even at the lower end of “elevated BP” spectrum, dilution of risk discrimination, and limited applicability in preventive medicine settings.

In this study, we sought to examine the consequences of the 2024 ESC BP classification in a preventive cohort in terms of prevalence and risk distribution. This cohort, part of a preventive medicine program conducted in a dedicated health assessment center in Paris, France, is composed of urban, highly educated individuals from higher socioeconomic backgrounds, actively engaged in their health, with facilitated access to care, a population expected to have an even lower burden of cardiovascular risk factors than the general French population sampled in the CONSTANCES cohort (12). Using data from participants undergoing comprehensive health assessments in this setting, we aimed to: (a) quantify the shift in BP category prevalence under the new ESC 2024 guidelines, and (b) characterize the clinical and biological profile of individuals newly reclassified as having “elevated” BP despite a previously “optimal” classification. Our hypothesis was that this reclassification of patients with BP <120/80 mmHg but DBP in the 70-79 mmHg range does not meaningfully distinguish a higher-risk group and may reflect an overly sensitive threshold of diastolic BP.

## Material & Methods

### Study Design and Ethical Approval

This cross-sectional study includes all adults who underwent a standardized, multidimensional health check-up at a dedicated preventive health center in Paris, France. For this analysis, we selected all participants assessed from November 2023 to May 2025 who had complete and validated office blood pressure (BP) measurements (n□=□1,394). All participants provided written informed consent for the use of their pseudonymized data for research purposes. The study protocol was reviewed and approved by an independent Institutional Review Board (Groupe Adene, IRB accreditation number: 0990-0279). Data handling and processing complied with the European General Data Protection Regulation (GDPR) and French MR-004 legal requirements.

### Blood Pressure Measurement Protocol

BP was measured in-office by trained healthcare staff using a validated OMRON EVOLV automatic oscillometric device (Omron Healthcare Co., Kyoto, Japan), in accordance with the ESC/ESH recommendations for automated BP monitoring (13). BP was measured according to a standardized protocol, detailed in Supplementary Method.

### Blood Pressure Classification

Participants were **categorized** using three guideline-based systems:

- **ESC 2024** (4): Non-elevated (<120/70 mmHg), Elevated (120–139 and/or 70–89 mmHg), Hypertension (≥140/90 mmHg or treatment)
- **ESC/ESH 2018** (5): Optimal (<120/80 mmHg), Normal (120–129/80–84 mmHg), High-Normal (130–139/85–89 mmHg), Hypertension (≥140/90 mmHg or treatment)

Analyses were first performed in the entire study population to describe BP category distributions across ESC/ESH2018 and ESC 2024 definitions. We then conducted targeted comparisons: (i) individuals reclassified from ESC/ESH2018 Optimal to ESC 2024 Elevated based solely on DBP ≥ 70 mmHg with SBP < 120 mmHg versus those who remained ESC/ESH2018 Optimal / ESC 2024 Non-elevated; (ii) ESC/ESH2018 Normal / ESC 2024 Elevated versus ESC/ESH2018 Optimal / ESC 2024 Elevated, to assess whether risk increases in the Normal/Elevated group where SBP is 120–129 mmHg, unlike the “diastolic-only” Optimal/Elevated group; (iii) ESC/ESH2018 Optimal versus all non-optimal participants, to highlight the broader discriminative capacity of the 2018 classification.

### Clinical, Lifestyle, and Biological Data

Other collected variables included demographic data (age, sex, body mass index, and educational level: primary, secondary, undergraduate, or graduate and above), lifestyle factors (smoking status as current, former, or never, with tobacco smoking pack-years, and alcohol consumption frequency and quantity), and self-reported health information (treatments, sleep apnea, diabetes, and other cardiovascular diseases). Biological data comprised a fasting lipid profile (total cholesterol, HDL, LDL, triglycerides), homocysteine, cystatin C, creatinine, estimated glomerular filtration rate (using the CKD-EPI equation) (14), and bone density markers for osteoporosis. Cardiovascular risk was estimated using the SCORE2 algorithm (15) for participants with complete data on age, sex, systolic blood pressure, smoking status, and total cholesterol. This was a cross-sectional analysis based on a single time-point assessment, with no follow-up data on cardiovascular events.

### Statistical Analysis

Descriptive statistics were used to summarize participant characteristics. Continuous variables were expressed as means□±□standard deviation or medians with interquartile range, and categorical variables as counts and percentages. Group comparisons used chiLsquare tests for categorical variables and linear regression (or logistic regression for binary outcomes) for continuous variables; ordinal regression was used across ordered BP categories. A two□sided p□<□0.05 was considered significant. All statistical analyses were performed using R software (version□4.4.2).

## Results

### Study Population

A total of 1,394 participants were included. The mean age was 49.9□±□12.1 years, and 921 (66.1L%) were men. Mean BMI was 24.2□±□3.99□kg/m². By BMI category, 63.0□% were normal weight, 29.0□% overweight, and 8.0□% obese. Educational level was high, with 72.3□% holding a graduate degree or higher. Daily alcohol consumption was reported by 121 (8.7%), 296 (21.2%) were former smokers, and 188 (13.5%) were current smokers with a mean packLyears of 10.0□±□12.6. Mean diastolic BP was 81.3□±□10.1LmmHg, and mean systolic BP was 124□±□14.2LmmHg. Hypertension treatment was reported by 4.8L% (**Table 1**). Baseline characteristics by ESC/ESH 2018 and ESC 2024 categories are presented in **Table S1**. As expected, individuals with higher BP categories were older, more often male, with higher BMI, and reported more comorbidities and treatment for hypertension, while those in the Optimal or Non-elevated categories had a more favorable profile.

**Table 1.**
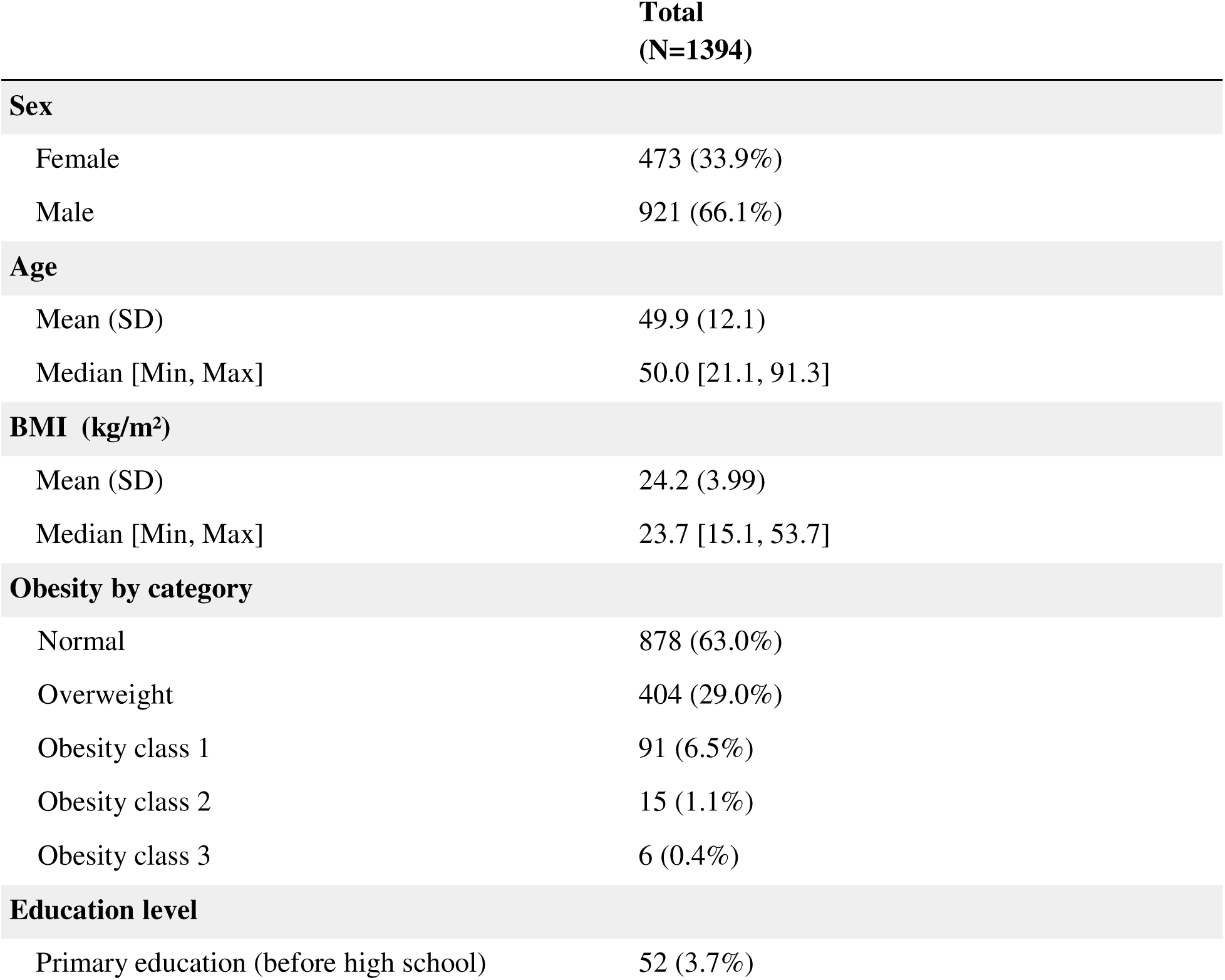
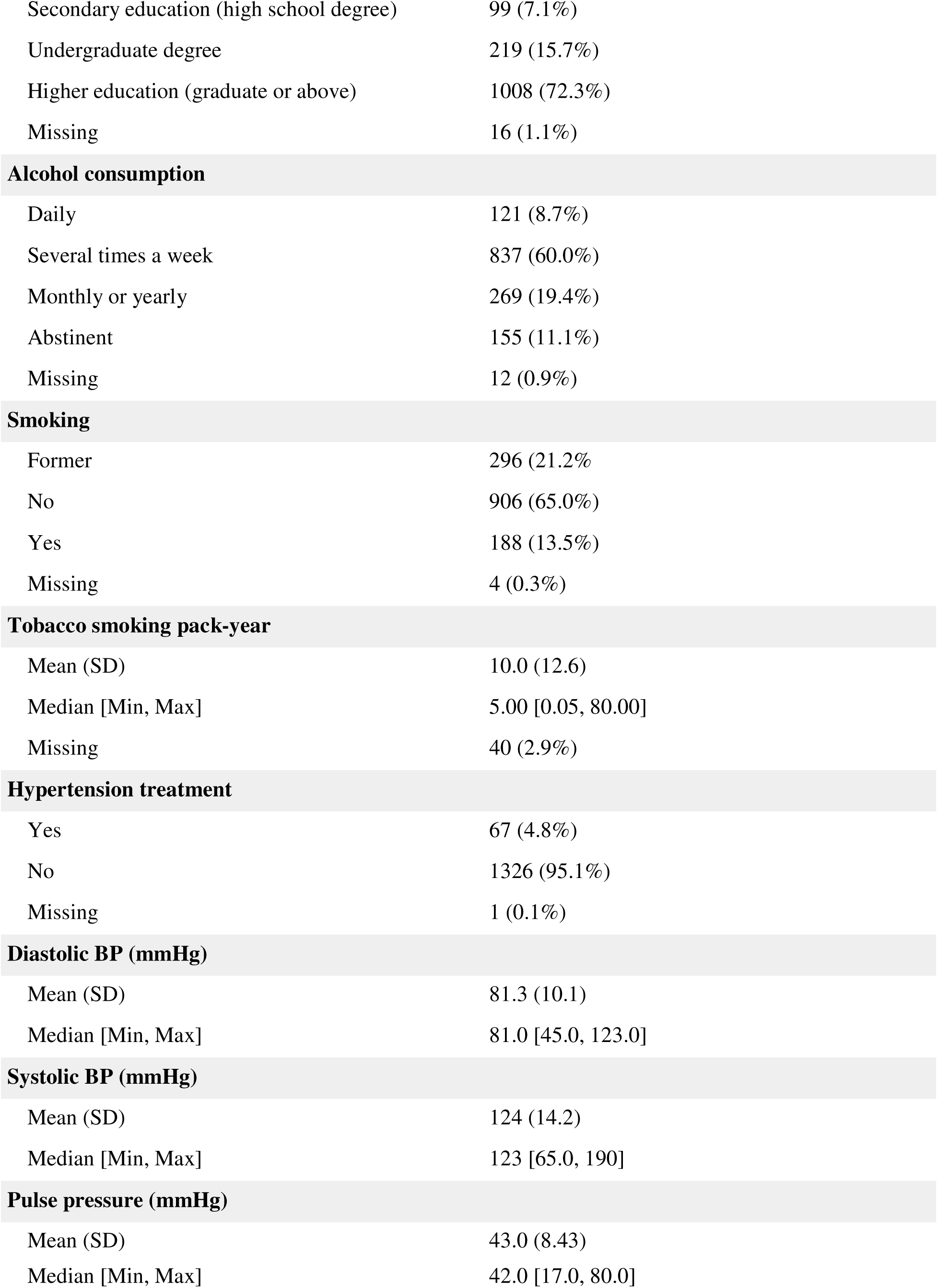
Baseline demographic, clinical, and lifestyle characteristics of the study population (N□=□1,394)

### Blood Pressure Classification

Under the ESC/ESH 2018 guidelines, 33.5% of participants had “Optimal” BP, 22.7% were “Normal,” 18.1% “High-normal,” and 25.8% were classified as hypertensive. With the ESC 2024 guidelines, only 10.0% remained in the “Non-elevated” category, while 64.2% were reclassified as having “Elevated” BP, and 25.8% remained hypertensive. **Table S2** summarizes the distribution of participants across these classification systems.

Figure 1 illustrates the shift in BP distribution between ESC/ESH 2018 and ESC 2024 classifications.

**Figure 1.**
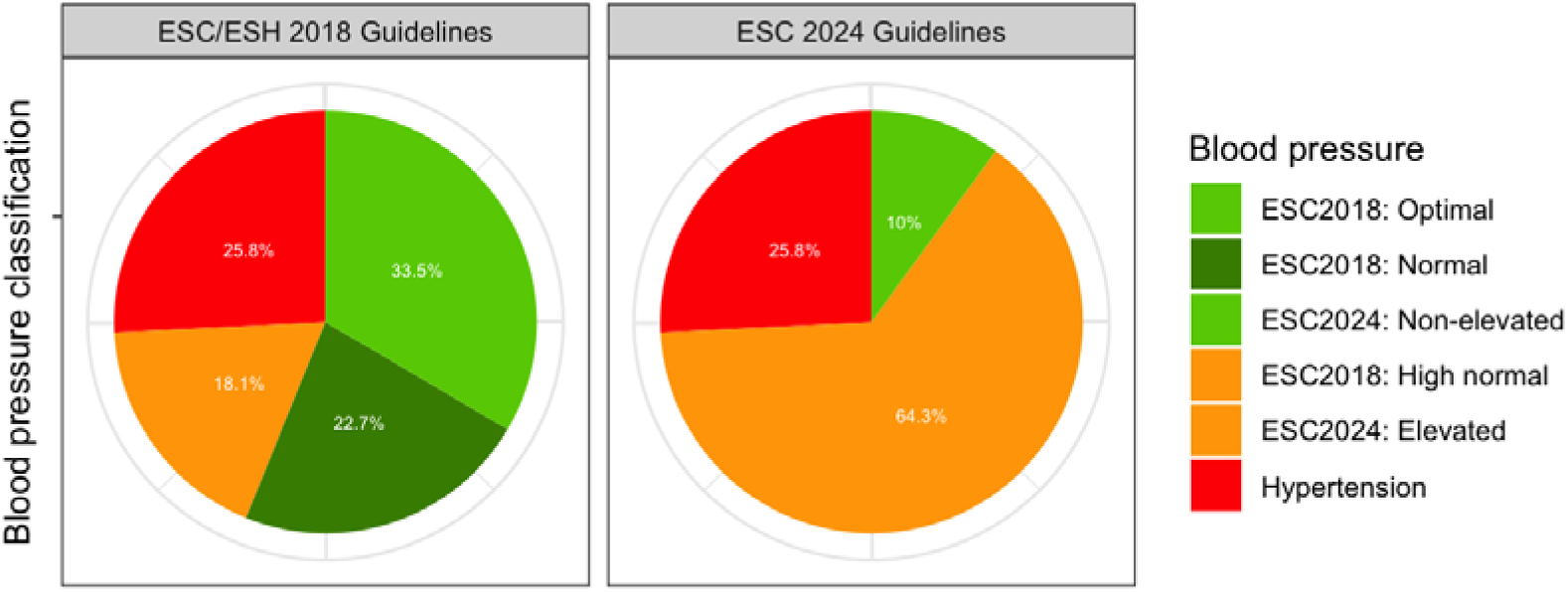
Distribution of blood pressure classification according to the ESC guidelines (2018 vs. 2024)

To better understand the dynamic introduced by the ESC 2024 update, we further examined cross-classifications between ESC/ESH 2018 and ESC 2024. As shown in Figure 2, a substantial proportion of participants initially classified as “Optimal” under ESC/ESH 2018 were reclassified as “Elevated” under ESC 2024.

**Figure 2.**
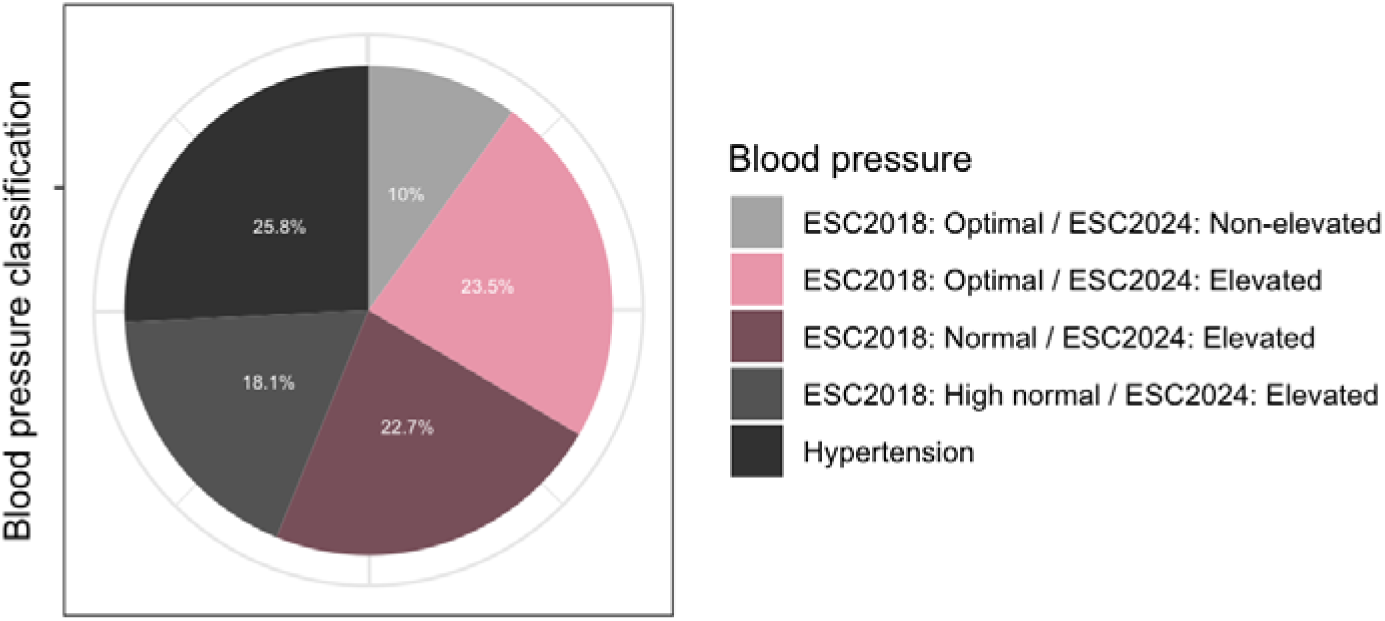
Distribution of blood pressure classification switch between ESC/ESH 2018 and ESC 2024 definitions.

### Comparison of Reclassified Groups

A subgroup of primary interest comprised individuals reclassified from ESC/ESH□2018: Optimal to ESCL2024: Elevated (n□=□328; 23.5%). This diastolicLonly reclassification (DBP□≥□70□mmHg with SBP□<□120□mmHg) was compared with participants who remained ESC/ESH□2018: Optimal / ESCL2024: Non□elevated (n□=□139; 10.0%).

As shown in **Table 3**, the groups were broadly similar for sex, BMI (including overweight/obesity grades), smoking exposure (pack-years), alcohol pattern, self-rated health, CVD history, diabetes, sleep apnea, and routine biomarkers. Age was the only non-BP variable that differed (45.5 vs. 42.7 years; p = 0.007). Hemodynamically, SBP was higher and pulse pressure (PP) lower in the reclassified group (both p < 0.001), consistent with a diastolic-driven shift and SBP < 120 mmHg. In regression analyses (binary and ordinal), age remained the only covariate associated with reclassification (**Table S3**). This association persisted after adjustment for SBP (p = 0.003), while additional adjustment for pulse pressure led to unstable estimates due to collinearity between SBP and PP.

**Table 3.**
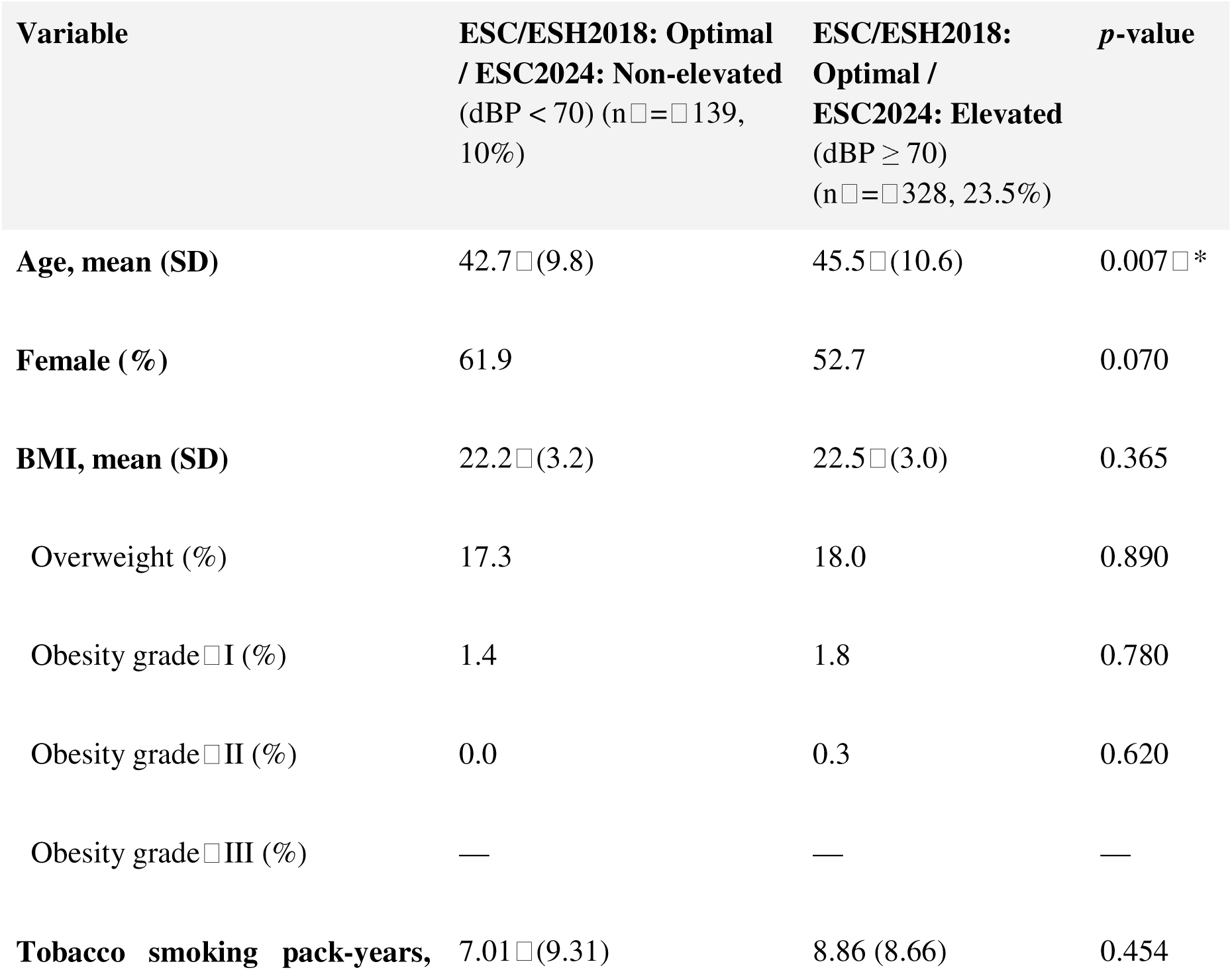
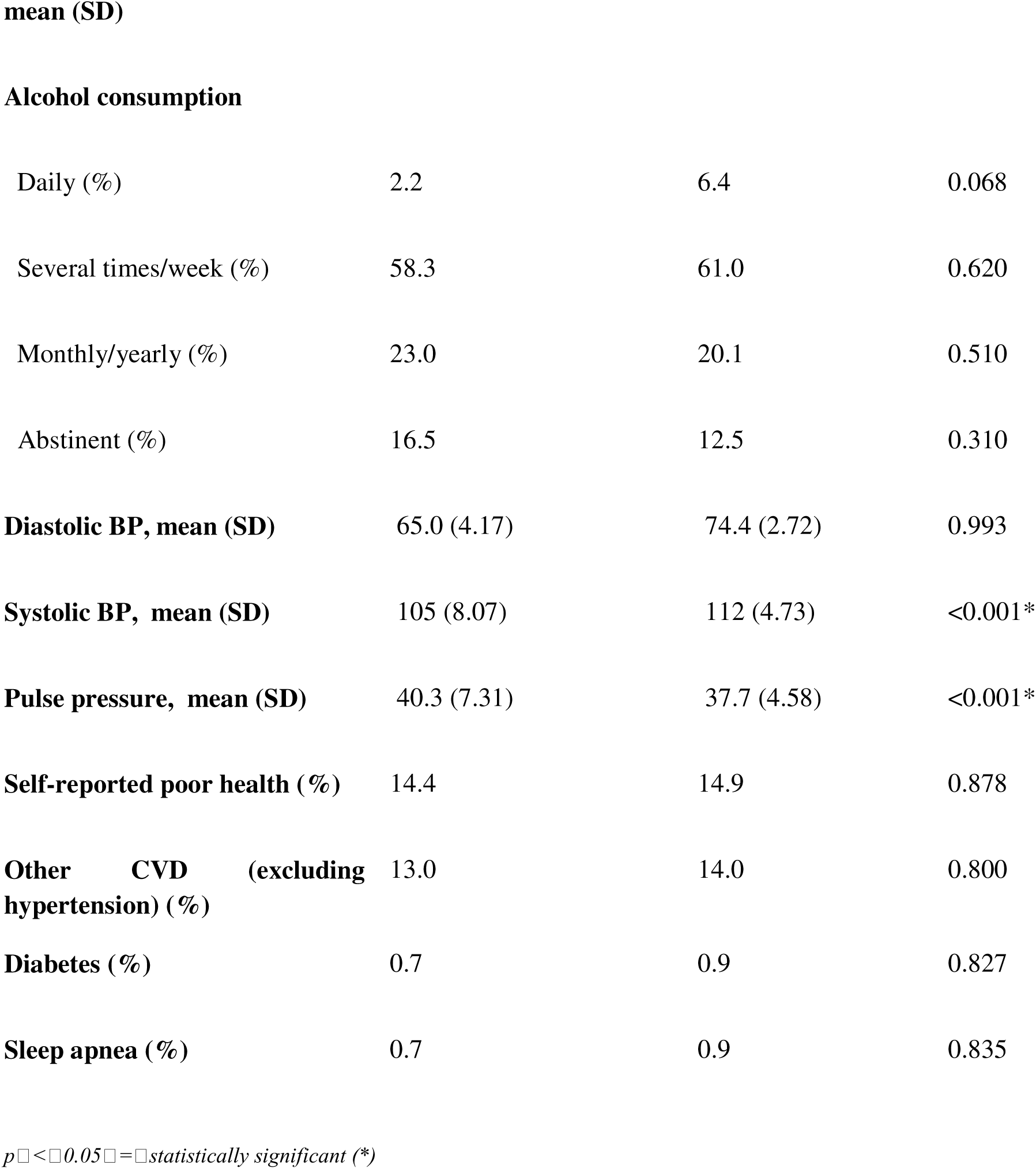
Comparison of clinical and demographic variables between “ESC/ESH 2018: Optimal / ESC2024: Non-elevated” and “ESC/ESH 2018: Optimal / ESC2024: Elevated” groups

| Variable | ESC/ESH2018: Optimal / ESC2024: Non-elevated (dBP < 70) ($n = 139$, 10%) | ESC/ESH2018: Optimal / ESC2024: Elevated (dBP $\geq 70$) ($n = 328$, 23.5%) | *p*-value |
| --- | --- | --- | --- |
| Age, mean (SD) | 42.7 (9.8) | 45.5 (10.6) | 0.007 * |
| Female (%) | 61.9 | 52.7 | 0.070 |
| BMI, mean (SD) | 22.2 (3.2) | 22.5 (3.0) | 0.365 |
| Overweight (%) | 17.3 | 18.0 | 0.890 |
| Obesity grade I (%) | 1.4 | 1.8 | 0.780 |
| Obesity grade II (%) | 0.0 | 0.3 | 0.620 |
| Obesity grade III (%) | — | — | — |
| Tobacco smoking pack-years, | 7.01 (9.31) | 8.86 (8.66) | 0.454 |
| **Alcohol consumption** |  |  |  |
| Daily (%) | 2.2 | 6.4 | 0.068 |
| Several times/week (%) | 58.3 | 61.0 | 0.620 |
| Monthly/yearly (%) | 23.0 | 20.1 | 0.510 |
| Abstinent (%) | 16.5 | 12.5 | 0.310 |
| **Diastolic BP, mean (SD)** | 65.0 (4.17) | 74.4 (2.72) | 0.993 |
| **Systolic BP, mean (SD)** | 105 (8.07) | 112 (4.73) | <0.001* |
| **Pulse pressure, mean (SD)** | 40.3 (7.31) | 37.7 (4.58) | <0.001* |
| **Self-reported poor health (%)** | 14.4 | 14.9 | 0.878 |
| **Other CVD (excluding hypertension) (%)** | 13.0 | 14.0 | 0.800 |
| **Diabetes (%)** | 0.7 | 0.9 | 0.827 |
| **Sleep apnea (%)** | 0.7 | 0.9 | 0.835 |

$p < 0.05$ = statistically significant (\*)

We next contrasted the two largest 2024 Elevated subsets: ESC/ESH2018: Normal / ESC2024: Elevated (n = 316; 22.7%) and ESC/ESH2018: Optimal / ESC2024: Elevated (n = 328; 23.5%).

As detailed in **Table 4,** in contrast with the comparisons of the two subpopulations of optimal BP (shown in table 3), the ESC/ESH2018: Normal/ESC□2024: Elevated subgroup showed a less favorable profile than ESC/ESH2018: Optimal/ ESCL2024: Elevated subgroup (older, fewer women, higher BMI, higher SBP/DBP, wider pulse pressure, and more sleep apnea; all p□≤□0.008).

**Table 4.** Comparison of “ESC/ESH2018: Normal / ESC2024: Elevated” vs. “ESC/ESH2018: Optimal / ESC2024: Elevated”.

| **Variable** | **ESC/ESH2018: Normal / ESC2024: Elevated (dBP $\geq 70$) (n = 316; 22.7%)** | **ESC/ESH2018: Optimal / ESC2024: Elevated (dBP $\geq 70$) (n = 328, 23.5%)** | **p-value** |
| --- | --- | --- | --- |
| **Age, mean (SD)** | 48.3 (11.2) | 45.5 (10.6) | 0.002* |
| **Female (%)** | 28.2 | 52.7 | <0.001* |
| **BMI, mean (SD)** | 24.1 (3.81) | 22.5 (3.0) | <0.001* |
| Overweight (%) | 25.6 | 18.0 | 0.064 |
| Obesity grade I (%) | 3.48 | 1.8 | 0.305 |
| Obesity grade II (%) | 1.27 | 0.3 | 0.974 |
| Obesity grade III (%) | 0.633 | — | — |
| **Tobacco smoking pack-years, mean (SD)** | 11.4 (17.7) | 8.86 (8.66) | 0.412 |
| **Alcohol consumption** |  |  |  |
| Daily (%) | 7.59 | 6.4 | 0.567 |
| Several times/week (%) | 64.2 | 61.0 | 0.772 |
| Monthly/yearly (%) | 17.4 | 20.1 | 0.819 |
| Abstinent (%) | 10.4 | 12.5 | 0.792 |
| **Diastolic BP, mean (SD)** | 79.3 (4.18) | 74.4 (2.72) | <0.001* |
| **Systolic BP, mean (SD)** | 122 (4.64) | 112 (4.73) | <0.001* |
| **Pulse pressure, mean (SD)** | 42.7 (6.62) | 37.7 (4.58) | <0.001* |
| **Self-reported poor health (%)** | 15.8 | 14.9 | 0.756 |
| **Other CVD (excluding hypertension) (%)** | 20.3 | 14.0 | 0.077 |
| **Diabetes (%)** | 2.2 | 0.9 | 0.200 |
| **Sleep apnea (%)** | 4.7 | 0.9 | 0.008 |

$p < 0.05$ = statistically
significant

Finally, for completeness, **Table S4** summarizes the broader contrast between ESC/ESH2018: Optimal and all Non-Optimal participants. As expected, Non-Optimal individuals were older, more often male, heavier, with higher BP values and greater comorbidity burden, while smoking exposure was similar.

### SCORE2 Risk Distribution

The SCORE2 cardiovascular risk score was used to further investigate differences across BP groups. A shown in Figure 3, SCORE2 values increased progressively from ESC/ESH2018: Optimal / ESC2024: Non-elevated to Hypertension (p < 0.001). No significant difference was observed between ESC/ESH2018: Optimal / ESC2024: Non-elevated and ESC/ESH2018: Optimal / ESC2024: Elevated (p = 0.524). By contrast, ESC/ESH2018: Normal / ESC2024: Elevated already showed significantly higher SCORE2 values compared with Optimal / Elevated (p = 0.017), supporting the greater clinical relevance of this subgroup.

**Figure 3.**
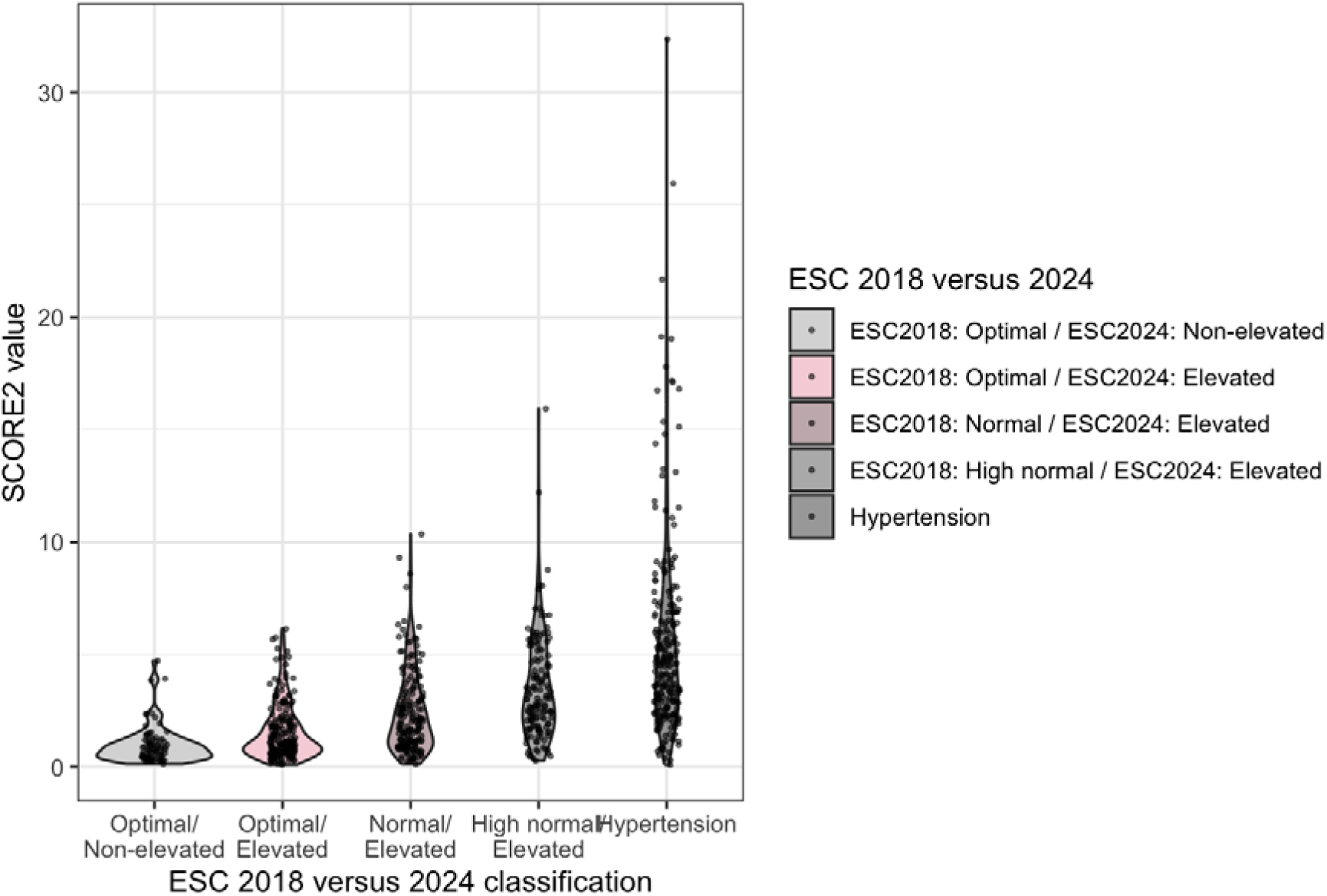
SCORE2 risk continuum across blood pressure categories.

## Discussion

In this study of 1,394 adults undergoing standardized preventive health evaluations, applying the 2024 ESC blood pressure classification markedly reshaped the distribution of BP categories. Overall, 23.5% of participants were reclassified from ESC/ESH2018: Optimal to ESC2024: Elevated, driven solely by diastolic blood pressure (DBP) ≥ 70 mmHg with systolic blood pressure (SBP) < 120 mmHg.These reclassified individuals accounted for nearly 70% of the 2018 Optimal BP subgroup. An additional 22.7% transitioned from ESC/ESH2018: Normal to ESC2024: Elevated, reflecting the combined effect of systolic and diastolic cutoffs.

Participants reclassified from Optimal to Elevated on the basis of DBP ≥ 70 mmHg displayed a very similar risk profile to those who remained Optimal/Non-elevated. Across clinical, demographic, and behavioral parameters, including BMI, smoking exposure, alcohol intake, self-rated health, comorbidities, and biomarkers, no meaningful differences were observed. The only variable significantly associated with reclassification was age (45.5 vs. 42.7 years; p = 0.007), likely due to a slightly higher mean SBP level in the reclassified group. Indeed, hemodynamically, this group showed higher DBP (**74.4 vs 65.0 mmHg**) and slightly higher SBP (112 vs 105 mmHg), both < 120 mmHg by definition, with narrower pulse pressure (37.7 vs 40.3 mmHg), consistent with the diastolic-driven reclassification. In a logistic model, the association between age and reclassification persisted after adjustment for SBP (p = 0.003), whereas adding pulse pressure produced unstable estimates consistent with collinearity between SBP and PP. Importantly, cardiovascular risk assessed by SCORE2 did not differ between the Optimal/Non-elevated and Optimal/Elevated groups (15), highlighting the limited discriminative power of this new DBP threshold in a low-risk preventive population.

By contrast, individuals reclassified from Normal to Elevated under the 2024 guidelines (n = 316; 22.7%) exhibited a less favorable profile compared with the Optimal/Elevated subgroup. They were older, more often male, and had higher BMI with a greater prevalence of overweight/obesity. Cardiometabolic comorbidities were also more frequent, including sleep apnea, while hemodynamic differences were striking: higher SBP, higher DBP, and wider pulse pressure (all p < 0.001). Unlike the Optimal/Elevated reclassification, these findings suggest that the Normal/Elevated group may indeed represent a higher-risk population, supporting the clinical legitimacy of this reclassification. Overall, this may argue in favor of the latest ACC/AHA guideline classification (Jones et al JACC 2025) which defines normal BP below 120/80 mmHg.Taken together, our data point to a more decisive role of SBP for risk stratification, and are compatible with frameworks that define normal BP below 120/80 mmHg, without presupposing a single optimal classification scheme.

Our findings echo those from the CONSTANCES cohort (11), where reducing the DBP threshold reclassified a substantial proportion of previously Optimal individuals without evidence of higher risk in the 6.5 year longitudinal follow-up of the cohort. Large-scale epidemiological studies consistently show a log-linear relationship between SBP and cardiovascular outcomes (6,16), while associations with DBP are weaker and less consistent (10,17). Moreover, very low DBP (< 70 mmHg) has been linked to adverse outcomes through impaired coronary perfusion, particularly in older individuals with atherosclerosis (8,18,19), supporting a U-shaped rather than strictly linear relationship.

A strength of this study is the detailed phenotyping of a preventive cohort, with standardized BP assessment and rich clinical/behavioral data. However, participants were highly educated, health-conscious, and low-risk, limiting generalizability to the broader population. ESC/ESH recommendations are partly designed to address underdiagnosis and undertreatment of hypertension in the general population, where up to half of hypertensive individuals remain unaware of their condition (20). The lowered DBP threshold may thus have greater utility in high-risk or less-well monitored settings than in preventive cohorts such as the present one. From a practical standpoint in primary prevention, a diastolic-only “elevated BP” label (DBP 70–79 mmHg with SBP < 120 mmHg) may warrant confirmation on repeated office or out-of-office measurements and a focus on lifestyle interventions rather than a change in medical status per se.The main limitation of this work is that it relies on cardiovascular risk scores and clinical/biological markers rather than adjudicated outcome events. Evaluating the true prognostic impact of this reclassification would require much larger cohorts and long-term follow-up with hard endpoints.

In this preventive medicine cohort, lowering the diastolic threshold in the 2024 ESC/ESH guidelines led to a large reclassification of Optimal individuals, but this “diastolic-only” group showed no evidence of higher clinical or biological risk compared with Non-elevated peers. By contrast, those reclassified from Normal to Elevated exhibited a less favorable risk profile, suggesting greater clinical relevance. These findings highlight the risk of overdiagnosis when applying the new DBP threshold in low-risk populations, while underscoring the importance of focusing reclassification efforts where they most strongly align with cardiovascular risk. Prospective outcome studies are needed to determine the true clinical impact of this change.

## Data Availability

All data produced in the present study are available upon reasonable request to the authors

## Reference

1. Forouzanfar MH, Afshin A, Alexander LT, Anderson HR, Bhutta ZA, Biryukov S, et al. Global, regional, and national comparative risk assessment of 79 behavioural, environmental and occupational, and metabolic risks or clusters of risks, 1990–2015: a systematic analysis for the Global Burden of Disease Study 2015. The Lancet. 2016 Oct;388(10053):1659–724.

2. Forouzanfar MH, Liu P, Roth GA, Ng M, Biryukov S, Marczak L, et al. Global Burden of Hypertension and Systolic Blood Pressure of at Least 110 to 115 mm Hg, 1990-2015. JAMA. 2017 Jan 10;317(2):165.

3. Kuneinen SM, Kautiainen H, Ekblad MO, Korhonen PE. Multifactorial prevention program for cardiovascular disease in primary care: hypertension status and effect on mortality. J Hum Hypertens. 2024 Feb 20;38(4):322–8.

4. McEvoy JW, McCarthy CP, Bruno RM, Brouwers S, Canavan MD, Ceconi C, et al. 2024 ESC Guidelines for the management of elevated blood pressure and hypertension. European Heart Journal. 2024 Oct 7;45(38):3912–4018.

5. Williams B, Mancia G, Spiering W, Agabiti Rosei E, Azizi M, Burnier M, et al. 2018 ESC/ESH Guidelines for the management of arterial hypertension. European Heart Journal. 2018 Sept 1;39(33):3021–104.

6. Age-specific relevance of usual blood pressure to vascular mortality: a meta-analysis of individual data for one million adults in 61 prospective studies. The Lancet. 2002 Dec;360(9349):1903–13.

7. Whelton SP, McEvoy JW, Shaw L, Psaty BM, Lima JAC, Budoff M, et al. Association of Normal Systolic Blood Pressure Level With Cardiovascular Disease in the Absence of Risk Factors. JAMA Cardiol. 2020 Sept 1;5(9):1011.

8. Anderson TerenceW. RE-EXAMINATION OF SOME OF THE FRAMINGHAM BLOOD-PRESSURE DATA. The Lancet. 1978 Nov;312(8100):1139–41.

9. Rahimi K, Bidel Z, Nazarzadeh M, Copland E, Canoy D, Ramakrishnan R, et al. Pharmacological blood pressure lowering for primary and secondary prevention of cardiovascular disease across different levels of blood pressure: an individual participant-level data meta-analysis. The Lancet. 2021 May;397(10285):1625–36.

10. McEvoy JW, Chen Y, Rawlings A, Hoogeveen RC, Ballantyne CM, Blumenthal RS, et al. Diastolic Blood Pressure, Subclinical Myocardial Damage, and Cardiac Events. Journal of the American College of Cardiology. 2016 Oct;68(16):1713–22.

11. Vidal-Petiot E, Kab S, Steg PG. New Definition of Elevated Blood Pressure in the 2024 ESC Guidelines: Increased Prevalence, Uncertain Evidence. Circulation. 2025 Feb 25;151(8):518–20.

12. Bauvin P, Benani A, Lepoittevin M, Sentilhes M, Bringer M, Lecheheb DA, et al. A Comprehensive Prospective Cohort in Preventive Medicine: Protocol and Profile of the First 1,000 Participants in a Health Screening Program [Internet]. Epidemiology; 2025 [cited 2025 Aug 5]. Available from: http://medrxiv.org/lookup/doi/10.1101/2025.07.07.25330615

13. Hakuo Takahashi. Validation of the Omron EVOLV (HEM-7600T-E) upper arm blood pressure monitor, in oscillometry mode, for self measurement in a general population, according to the European Society of Hypertension International Protocol revision 2010. Available from: http://www.dableducational.org/Publications/2016/ESH-IP%202010%20Validation%20of%20HEM-7600T-E.pdf

14. Lu S, Robyak K, Zhu Y. The CKD-EPI 2021 Equation and Other Creatinine-Based Race-Independent eGFR Equations in Chronic Kidney Disease Diagnosis and Staging. The Journal of Applied Laboratory Medicine. 2023 Sept 7;8(5):952–61.

15. SCORE2 working group and ESC Cardiovascular risk collaboration, Hageman S, Pennells L, Ojeda F, Kaptoge S, Kuulasmaa K, et al. SCORE2 risk prediction algorithms: new models to estimate 10-year risk of cardiovascular disease in Europe. European Heart Journal. 2021 July 1;42(25):2439–54.

16. Ettehad D, Emdin CA, Kiran A, Anderson SG, Callender T, Emberson J, et al. Blood pressure lowering for prevention of cardiovascular disease and death: a systematic review and meta-analysis. The Lancet. 2016 Mar;387(10022):957–67.

17. Kjeldsen SE. Hypertension and cardiovascular risk: General aspects. Pharmacological Research. 2018 Mar;129:95–9.

18. Franklin SS, Gustin W, Wong ND, Larson MG, Weber MA, Kannel WB, et al. Hemodynamic Patterns of Age-Related Changes in Blood Pressure: The Framingham Heart Study. Circulation. 1997 July;96(1):308–15.

19. Rahman F, Al Rifai M, Blaha MJ, Nasir K, Budoff MJ, Psaty BM, et al. Relation of Diastolic Blood Pressure and Coronary Artery Calcium to Coronary Events and Outcomes (From the Multi-Ethnic Study of Atherosclerosis). Am J Cardiol. 2017 Nov 15;120(10):1797–803.

20. Mills KT, Bundy JD, Kelly TN, Reed JE, Kearney PM, Reynolds K, et al. Global Disparities of Hypertension Prevalence and Control: A Systematic Analysis of Population-Based Studies From 90 Countries. Circulation. 2016 Aug 9;134(6):441–50.

