## Supplemental File for "Comparison of cardiovascular risk in individuals with normal vs isolated elevated diastolic blood pressure"

**Supplementary File**

**Supplementary Methods – Blood Pressure Measurement Protocol**

Blood pressure was measured in a quiet, standardized environment with controlled room temperature (> 24.5 °C). Participants rested for at least 15 minutes before measurement. Measurements were taken with patients seated, using a cuff appropriate to arm size, the arm supported at heart level, and feet flat on the floor. Participants were instructed to refrain from smoking, exercising, eating, or consuming caffeine for at least 30 minutes prior to measurement.

**Table S1**. **A.** Baseline characteristics by ESC/ESH 2018 categories, **B**.Baseline characteristics by ESC 2024 categories
A.

| *ESC/ESH 2018* | **Optimal**  **(N=467)** | **Normal**  **(N=316)** | **High normal**  **(N=252)** | **Hypertension**  **(N=359)** |
| --- | --- | --- | --- | --- |
| **Sex** |  |  |  |  |
| Female | 259 (55.5%) | 89 (28.2%) | 53 (21.0%) | 72 (20.1%) |
| Male | 208 (44.5%) | 227 (71.8%) | 199 (79.0%) | 287 (79.9%) |
| **Age** |  |  |  |  |
| Mean (SD) | 44.7 (10.4) | 48.3 (11.2) | 52.0 (11.0) | 56.6 (11.9) |
| Median [Min, Max] | 43.9 [21.1, 81.6] | 48.5 [22.7, 78.3] | 52.7 [21.4, 80.3] | 57.0 [23.0, 91.3] |
| **BMI (kg/m²)** |  |  |  |  |
| Mean (SD) | 22.4 (3.04) | 24.1 (3.81) | 25.0 (3.66) | 26.1 (4.40) |
| Median [Min, Max] | 22.0 [15.1, 38.6] | 23.5 [16.4, 48.9] | 24.8 [17.7, 46.1] | 25.4 [16.8, 53.7] |
| **Obesity by category** |  |  |  |  |
| Normal | 371 (79.4%) | 218 (69.0%) | 130 (51.6%) | 159 (44.3%) |
| Overweight | 87 (18.6%) | 81 (25.6%) | 99 (39.3%) | 137 (38.2%) |
| Obesity class 1 | 8 (1.7%) | 11 (3.5%) | 21 (8.3%) | 51 (14.2%) |
| Obesity class 2 | 1 (0.2%) | 4 (1.3%) | 1 (0.4%) | 9 (2.5%) |
| Obesity class 3 | 0 (0%) | 2 (0.6%) | 1 (0.4%) | 3 (0.8%) |
| **Education level** |  |  |  |  |
| Primary education (before high school) | 14 (3.0%) | 9 (2.8%) | 6 (2.4%) | 23 (6.4%) |
| Secondary education (high school degree) | 33 (7.1%) | 16 (5.1%) | 14 (5.6%) | 36 (10.0%) |
| Undergraduate degree | 71 (15.2%) | 47 (14.9%) | 42 (16.7%) | 59 (16.4%) |
| Higher education (graduate or above) | 345 (73.9%) | 242 (76.6%) | 184 (73.0%) | 237 (66.0%) |
| Missing | 4 (0.9%) | 2 (0.6%) | 6 (2.4%) | 4 (1.1%) |
| **Alcohol consumption** |  |  |  |  |
| Daily | 24 (5.1%) | 24 (7.6%) | 24 (9.5%) | 49 (13.6%) |
| Several times a week | 301 (64.5%) | 203 (64.2%) | 147 (58.3%) | 186 (51.8%) |
| Monthly or yearly | 88 (18.8%) | 55 (17.4%) | 51 (20.2%) | 75 (20.9%) |
| Abstinent | 51 (10.9%) | 33 (10.4%) | 26 (10.3%) | 45 (12.5%) |
| Missing | 3 (0.6%) | 1 (0.3%) | 4 (1.6%) | 4 (1.1%) |
| **Smoking** |  |  |  |  |
| Former | 83 (17.8%) | 59 (18.7%) | 65 (25.8%) | 89 (24.8%) |
| No | 306 (65.5%) | 218 (69.0%) | 153 (60.7%) | 229 (63.8%) |
| Yes | 77 (16.5%) | 37 (11.7%) | 33 (13.1%) | 41 (11.4%) |
| Missing | 1 (0.2%) | 2 (0.6%) | 1 (0.4%) | 0 (0%) |
| **Paquets-Années** |  |  |  |  |
| Mean (SD) | 8.33 (8.83) | 11.4 (17.7) | 8.14 (8.79) | 13.2 (14.6) |
| Median [Min, Max] | 5.00 [0.150, 40.0] | 3.88 [0.0500, 80.0] | 3.57 [0.600, 30.0] | 9.50 [0.350, 54.0] |
| Missing | 401 (85.9%) | 282 (89.2%) | 223 (88.5%) | 321 (89.4%) |
| **Diastolic BP (mmHg)** |  |  |  |  |
| Mean (SD) | 71.6 (5.37) | 79.3 (4.18) | 84.9 (4.03) | 93.0 (7.73) |
| Median [Min, Max] | 73.0 [45.0, 79.0] | 80.0 [62.0, 84.0] | 86.0 [62.0, 89.0] | 92.0 [69.0, 123] |
| **Systolic BP (mmHg)** |  |  |  |  |
| Mean (SD) | 110 (6.70) | 122 (4.64) | 130 (5.44) | 140 (11.8) |
| Median [Min, Max] | 111 [65.0, 119] | 122 [103, 129] | 131 [113, 139] | 140 [104, 190] |
| **Pulse pressure (mmHg)** |  |  |  |  |
| Mean (SD) | 38.5 (5.65) | 42.7 (6.62) | 45.3 (7.55) | 47.4 (10.3) |
| Median [Min, Max] | 39.0 [18.0, 65.0] | 43.0 [23.0, 63.0] | 46.0 [26.0, 72.0] | 46.0 [17.0, 80.0] |
| **Hypertension treatment** |  |  |  |  |
| Yes | 0 (0%) | 0 (0%) | 0 (0%) | 67 (18.7%) |
| No | 467 (100%) | 316 (100%) | 252 (100%) | 291 (81.1%) |
| Missing | 0 (0%) | 0 (0%) | 0 (0%) | 1 (0.3%) |
| **Declared hypertension** |  |  |  |  |
| No | 463 (99.1%) | 309 (97.8%) | 244 (96.8%) | 276 (76.9%) |
| Yes | 3 (0.6%) | 6 (1.9%) | 4 (1.6%) | 82 (22.8%) |
| Missing | 1 (0.2%) | 1 (0.3%) | 4 (1.6%) | 1 (0.3%) |

**B.**

| *ESC 2024* | **Non-elevated**  **(N=139)** | **Elevated**  **(N=896)** | **Hypertension**  **(N=359)** |
| --- | --- | --- | --- |
| **Sex** |  |  |  |
| Female | 86 (61.9%) | 315 (35.2%) | 72 (20.1%) |
| Male | 53 (38.1%) | 581 (64.8%) | 287 (79.9%) |
| **Age** |  |  |  |
| Mean (SD) | 42.7 (9.78) | 48.3 (11.2) | 56.6 (11.9) |
| Median [Min, Max] | 41.2 [22.3, 70.6] | 48.4 [21.1, 81.6] | 57.0 [23.0, 91.3] |
| **BMI (kg/m²)** |  |  |  |
| Mean (SD) | 22.2 (3.19) | 23.8 (3.63) | 26.1 (4.40) |
| Median [Min, Max] | 21.8 [15.7, 38.6] | 23.4 [15.1, 48.9] | 25.4 [16.8, 53.7] |
| **Obesity by category** |  |  |  |
| Normal | 114 (82.0%) | 605 (67.5%) | 159 (44.3%) |
| Overweight | 23 (16.5%) | 244 (27.2%) | 137 (38.2%) |
| Obesity class 1 | 1 (0.7%) | 39 (4.4%) | 51 (14.2%) |
| Obesity class 2 | 1 (0.7%) | 5 (0.6%) | 9 (2.5%) |
| Obesity class 3 | 0 (0%) | 3 (0.3%) | 3 (0.8%) |
| **Education level** |  |  |  |
| Primary education (before high school) | 6 (4.3%) | 23 (2.6%) | 23 (6.4%) |
| Secondary education (high school degree) | 9 (6.5%) | 54 (6.0%) | 36 (10.0%) |
| Undergraduate degree | 15 (10.8%) | 145 (16.2%) | 59 (16.4%) |
| Higher education (graduate or above) | 109 (78.4%) | 662 (73.9%) | 237 (66.0%) |
| Missing | 0 (0%) | 12 (1.3%) | 4 (1.1%) |
| **Alcohol consumption** |  |  |  |
| Daily | 3 (2.2%) | 69 (7.7%) | 49 (13.6%) |
| Several times a week | 88 (63.3%) | 563 (62.8%) | 186 (51.8%) |
| Monthly or yearly | 29 (20.9%) | 165 (18.4%) | 75 (20.9%) |
| Abstinent | 19 (13.7%) | 91 (10.2%) | 45 (12.5%) |
| Missing | 0 (0%) | 8 (0.9%) | 4 (1.1%) |
| **Smoking** |  |  |  |
| Former | 33 (23.7%) | 174 (19.4%) | 89 (24.8%) |
| No | 82 (59.0%) | 595 (66.4%) | 229 (63.8%) |
| Yes | 23 (16.5%) | 124 (13.8%) | 41 (11.4%) |
| Missing | 1 (0.7%) | 3 (0.3%) | 0 (0%) |
| **Paquets-Années** |  |  |  |
| Mean (SD) | 7.09 (9.31) | 9.45 (12.2) | 13.2 (14.6) |
| Median [Min, Max] | 3.89 [0.500, 40.0] | 5.00 [0.0500, 80.0] | 9.50 [0.350, 54.0] |
| Missing | 119 (85.6%) | 787 (87.8%) | 321 (89.4%) |
| **Diastolic BP (mmHg)** |  |  |  |
| Mean (SD) | 65.0 (4.17) | 79.1 (5.55) | 93.0 (7.73) |
| Median [Min, Max] | 66.0 [45.0, 69.0] | 79.0 [62.0, 89.0] | 92.0 [69.0, 123] |
| **Systolic BP (mmHg)** |  |  |  |
| Mean (SD) | 105 (8.07) | 121 (8.76) | 140 (11.8) |
| Median [Min, Max] | 106 [65.0, 119] | 121 [97.0, 139] | 140 [104, 190] |
| **Pulse pressure (mmHg)** |  |  |  |
| Mean (SD) | 40.3 (7.31) | 41.6 (6.99) | 47.4 (10.3) |
| Median [Min, Max] | 41.0 [18.0, 65.0] | 41.0 [21.0, 72.0] | 46.0 [17.0, 80.0] |
| **Hypertension treatment** |  |  |  |
| Yes | 0 (0%) | 0 (0%) | 67 (18.7%) |
| No | 139 (100%) | 896 (100%) | 291 (81.1%) |
| Missing | 0 (0%) | 0 (0%) | 1 (0.3%) |
| **Declared hypertension** |  |  |  |
| No | 138 (99.3%) | 878 (98.0%) | 276 (76.9%) |
| Yes | 1 (0.7%) | 12 (1.3%) | 82 (22.8%) |
| Missing | 0 (0%) | 6 (0.7%) | 1 (0.3%) |

**Table S2.** Blood pressure classification of participants according to ESC/ESH 2018 and ESC 2024 guidelines

|  | **Total**  **(N=1394)** |
| --- | --- |
| **ESC/ESH 2018 Guidelines** |  |
| Optimal <120/80 | 467 (33.5%) |
| Normal <120-129/80-84 | 316 (22.7%) |
| High normal <130–139/85–89 | 252 (18.1%) |
| Hypertension ≥140/90 | 359 (25.8%) |
| **ESC 2024 Guidelines** |  |
| Non-elevated <120/70 | 139 (10.0%) |
| Elevated <139/89 | 896 (64.2%) |
| Hypertension ≥140/90 | 359 (25.8%) |

**Table S3**. Baseline demographic and clinical characteristics across blood pressure categories (ESC/ESH 2018 vs ESC 2024 definitions)

|  | **ESC/ESH2018: Optimal / ESC2024: Non-elevated**  **(N=139, 10%)** | **ESC/ESH2018: Optimal / ESC2024: Elevated**  **(N=328, 23.5%)** | **ESC/ESH2018: Normal / ESC2024: Elevated**  **(N=316, 22.7%)** | **ESC/ESH2018: High normal / ESC2024: Elevated**  **(N=252,18.1%)** | **Hypertension**  **(N=359, 25.8%)** | **Overall**  **(N=1394)** |
| --- | --- | --- | --- | --- | --- | --- |
| **Sex** |  |  |  |  |  |  |
| Female | 86 (61.9%) | 173 (52.7%) | 89 (28.2%) | 53 (21.0%) | 72 (20.1%) | 473 (33.9%) |
| Male | 53 (38.1%) | 155 (47.3%) | 227 (71.8%) | 199 (79.0%) | 287 (79.9%) | 921 (66.1%) |
| **Age** |  |  |  |  |  |  |
| Mean (SD) | 42.7 (9.78) | 45.5 (10.6) | 48.3 (11.2) | 52.0 (11.0) | 56.6 (11.9) | 49.9 (12.1) |
| Median [Min, Max] | 41.2 [22.3, 70.6] | 44.8 [21.1, 81.6] | 48.5 [22.7, 78.3] | 52.7 [21.4, 80.3] | 57.0 [23.0, 91.3] | 50.0 [21.1, 91.3] |
| **BMI (kg/m²)** |  |  |  |  |  |  |
| Mean (SD) | 22.2 (3.19) | 22.5 (2.98) | 24.1 (3.81) | 25.0 (3.66) | 26.1 (4.40) | 24.2 (3.99) |
| Median [Min, Max] | 21.8 [15.7, 38.6] | 22.1 [15.1, 32.3] | 23.5 [16.4, 48.9] | 24.8 [17.7, 46.1] | 25.4 [16.8, 53.7] | 23.7 [15.1, 53.7] |
| **Smoking** |  |  |  |  |  |  |
| Yes | 23 (16.5%) | 54 (16.5%) | 37 (11.7%) | 33 (13.1%) | 41 (11.4%) | 188 (13.5%) |
| No | 115 (82.7%) | 274 (83.5%) | 277 (87.7%) | 218 (86.5%) | 318 (88.6%) | 1202 (86.2%) |
| Missing | 1 (0.7%) | 0 (0%) | 2 (0.6%) | 1 (0.4%) | 0 (0%) | 4 (0.3%) |
| **Tobacco smoking pack-years** |  |  |  |  |  |  |
| Mean (SD) | 7.09 (9.31) | 8.86 (8.66) | 11.4 (17.7) | 8.14 (8.79) | 13.2 (14.6) | 10.0 (12.6) |
| Median [Min, Max] | 3.89 [0.500, 40.0] | 5.88 [0.150, 34.0] | 3.88 [0.0500, 80.0] | 3.57 [0.600, 30.0] | 9.50 [0.350, 54.0] | 5.00 [0.0500, 80.0] |
| Missing | 119 (85.6%) | 282 (86.0%) | 282 (89.2%) | 223 (88.5%) | 321 (89.4%) | 1227 (88.0%) |
| **Declare low health** |  |  |  |  |  |  |
| Yes | 20 (14.4%) | 49 (14.9%) | 50 (15.8%) | 39 (15.5%) | 101 (28.1%) | 259 (18.6%) |
| No | 119 (85.6%) | 279 (85.1%) | 266 (84.2%) | 213 (84.5%) | 258 (71.9%) | 1135 (81.4%) |
| **Alcohol** |  |  |  |  |  |  |
| Yes | 3 (2.2%) | 21 (6.4%) | 24 (7.6%) | 24 (9.5%) | 49 (13.6%) | 121 (8.7%) |
| No | 136 (97.8%) | 304 (92.7%) | 291 (92.1%) | 224 (88.9%) | 306 (85.2%) | 1261 (90.5%) |
| Missing | 0 (0%) | 3 (0.9%) | 1 (0.3%) | 4 (1.6%) | 4 (1.1%) | 12 (0.9%) |
| **Diastolic BP (mmHg)** |  |  |  |  |  |  |
| Mean (SD) | 65.0 (4.17) | 74.4 (2.72) | 79.3 (4.18) | 84.9 (4.03) | 93.0 (7.73) | 81.3 (10.1) |
| Median [Min, Max] | 66.0 [45.0, 69.0] | 74.0 [70.0, 79.0] | 80.0 [62.0, 84.0] | 86.0 [62.0, 89.0] | 92.0 [69.0, 123] | 81.0 [45.0, 123] |
| **Systolic BP (mmHg)** |  |  |  |  |  |  |
| Mean (SD) | 105 (8.07) | 112 (4.73) | 122 (4.64) | 130 (5.44) | 140 (11.8) | 124 (14.2) |
| Median [Min, Max] | 106 [65.0, 119] | 113 [97.0, 119] | 122 [103, 129] | 131 [113, 139] | 140 [104, 190] | 123 [65.0, 190] |
| **Pulse pressure (mmHg)** |  |  |  |  |  |  |
| Mean (SD) | 40.3 (7.31) | 37.7 (4.58) | 42.7 (6.62) | 45.3 (7.55) | 47.4 (10.3) | 43.0 (8.43) |
| Median [Min, Max] | 41.0 [18.0, 65.0] | 38.0 [21.0, 49.0] | 43.0 [23.0, 63.0] | 46.0 [26.0, 72.0] | 46.0 [17.0, 80.0] | 42.0 [17.0, 80.0] |
| **History CVD** |  |  |  |  |  |  |
| Yes | 19 (13.7%) | 49 (14.9%) | 64 (20.3%) | 48 (19.0%) | 131 (36.5%) | 311 (22.3%) |
| No | 120 (86.3%) | 278 (84.8%) | 251 (79.4%) | 200 (79.4%) | 227 (63.2%) | 1076 (77.2%) |
| Missing | 0 (0%) | 1 (0.3%) | 1 (0.3%) | 4 (1.6%) | 1 (0.3%) | 7 (0.5%) |
| **Measured obesity** |  |  |  |  |  |  |
| No | 137 (98.6%) | 321 (97.9%) | 299 (94.6%) | 229 (90.9%) | 296 (82.5%) | 1282 (92.0%) |
| Yes | 2 (1.4%) | 7 (2.1%) | 17 (5.4%) | 23 (9.1%) | 63 (17.5%) | 112 (8.0%) |
| **Diagnosed diabetes** |  |  |  |  |  |  |
| No | 134 (96.4%) | 312 (95.1%) | 299 (94.6%) | 236 (93.7%) | 334 (93.0%) | 1315 (94.3%) |
| Yes | 1 (0.7%) | 3 (0.9%) | 7 (2.2%) | 3 (1.2%) | 15 (4.2%) | 29 (2.1%) |
| Missing | 4 (2.9%) | 13 (4.0%) | 10 (3.2%) | 13 (5.2%) | 10 (2.8%) | 50 (3.6%) |
| **Measured hyperhomocysteinemia** |  |  |  |  |  |  |
| No | 74 (53.2%) | 187 (57.0%) | 209 (66.1%) | 171 (67.9%) | 232 (64.6%) | 873 (62.6%) |
| Yes | 61 (43.9%) | 128 (39.0%) | 98 (31.0%) | 68 (27.0%) | 116 (32.3%) | 471 (33.8%) |
| Missing | 4 (2.9%) | 13 (4.0%) | 9 (2.8%) | 13 (5.2%) | 11 (3.1%) | 50 (3.6%) |
| **Measured osteoporosis** |  |  |  |  |  |  |
| No | 120 (86.3%) | 285 (86.9%) | 254 (80.4%) | 217 (86.1%) | 288 (80.2%) | 1164 (83.5%) |
| Yes | 3 (2.2%) | 2 (0.6%) | 5 (1.6%) | 2 (0.8%) | 14 (3.9%) | 26 (1.9%) |
| Missing | 16 (11.5%) | 41 (12.5%) | 57 (18.0%) | 33 (13.1%) | 57 (15.9%) | 204 (14.6%) |
| **Measured renal deficiency** |  |  |  |  |  |  |
| No | 134 (96.4%) | 312 (95.1%) | 301 (95.3%) | 235 (93.3%) | 339 (94.4%) | 1321 (94.8%) |
| Yes | 0 (0%) | 3 (0.9%) | 5 (1.6%) | 2 (0.8%) | 9 (2.5%) | 19 (1.4%) |
| Missing | 5 (3.6%) | 13 (4.0%) | 10 (3.2%) | 15 (6.0%) | 11 (3.1%) | 54 (3.9%) |
| **Screened sleep apnea** |  |  |  |  |  |  |
| No | 138 (99.3%) | 325 (99.1%) | 301 (95.3%) | 239 (94.8%) | 305 (85.0%) | 1308 (93.8%) |
| Yes | 1 (0.7%) | 3 (0.9%) | 15 (4.7%) | 13 (5.2%) | 54 (15.0%) | 86 (6.2%) |
| **History stroke** |  |  |  |  |  |  |
| No | 134 (96.4%) | 315 (96.0%) | 302 (95.6%) | 233 (92.5%) | 330 (91.9%) | 1314 (94.3%) |
| Yes | 2 (1.4%) | 11 (3.4%) | 10 (3.2%) | 14 (5.6%) | 27 (7.5%) | 64 (4.6%) |
| Missing | 3 (2.2%) | 2 (0.6%) | 4 (1.3%) | 5 (2.0%) | 2 (0.6%) | 16 (1.1%) |
| **History heart rhythm disorder** |  |  |  |  |  |  |
| No | 136 (97.8%) | 311 (94.8%) | 299 (94.6%) | 237 (94.0%) | 326 (90.8%) | 1309 (93.9%) |
| Yes | 3 (2.2%) | 17 (5.2%) | 17 (5.4%) | 15 (6.0%) | 33 (9.2%) | 85 (6.1%) |
| **History cardiac insufficiency** |  |  |  |  |  |  |
| No | 138 (99.3%) | 327 (99.7%) | 314 (99.4%) | 251 (99.6%) | 355 (98.9%) | 1385 (99.4%) |
| Yes | 1 (0.7%) | 1 (0.3%) | 2 (0.6%) | 1 (0.4%) | 4 (1.1%) | 9 (0.6%) |
| **History heart murmur** |  |  |  |  |  |  |
| No | 136 (97.8%) | 325 (99.1%) | 305 (96.5%) | 251 (99.6%) | 345 (96.1%) | 1362 (97.7%) |
| Yes | 3 (2.2%) | 3 (0.9%) | 11 (3.5%) | 1 (0.4%) | 14 (3.9%) | 32 (2.3%) |
| **History myocardial infarction** |  |  |  |  |  |  |
| No | 137 (98.6%) | 326 (99.4%) | 313 (99.1%) | 249 (98.8%) | 350 (97.5%) | 1375 (98.6%) |
| Yes | 2 (1.4%) | 2 (0.6%) | 3 (0.9%) | 3 (1.2%) | 9 (2.5%) | 19 (1.4%) |
| **History phlebitis** |  |  |  |  |  |  |
| No | 136 (97.8%) | 326 (99.4%) | 310 (98.1%) | 249 (98.8%) | 350 (97.5%) | 1371 (98.4%) |
| Yes | 3 (2.2%) | 2 (0.6%) | 6 (1.9%) | 3 (1.2%) | 9 (2.5%) | 23 (1.7%) |
| **History venous disorder** |  |  |  |  |  |  |
| No | 138 (99.3%) | 327 (99.7%) | 313 (99.1%) | 251 (99.6%) | 355 (98.9%) | 1384 (99.3%) |
| Yes | 1 (0.7%) | 1 (0.3%) | 3 (0.9%) | 1 (0.4%) | 4 (1.1%) | 10 (0.7%) |
| **History tachycardia** |  |  |  |  |  |  |
| No | 136 (97.8%) | 323 (98.5%) | 309 (97.8%) | 249 (98.8%) | 351 (97.8%) | 1368 (98.1%) |
| Yes | 3 (2.2%) | 4 (1.2%) | 7 (2.2%) | 3 (1.2%) | 8 (2.2%) | 25 (1.8%) |
| Missing | 0 (0%) | 1 (0.3%) | 0 (0%) | 0 (0%) | 0 (0%) | 1 (0.1%) |
| **History bradycardia** |  |  |  |  |  |  |
| No | 139 (100%) | 326 (99.4%) | 315 (99.7%) | 251 (99.6%) | 356 (99.2%) | 1387 (99.5%) |
| Yes | 0 (0%) | 1 (0.3%) | 1 (0.3%) | 1 (0.4%) | 3 (0.8%) | 6 (0.4%) |
| Missing | 0 (0%) | 1 (0.3%) | 0 (0%) | 0 (0%) | 0 (0%) | 1 (0.1%) |

**Table S4.** Comparison of ESC/ESH2018 Optimal vs. all non-optimal participants (n = 1,394)

| **Variable** | **ESC/ESH2018 Optimal** (n=467, 33.5%) | **Non-Optimal** (n=927, 66.5%) | **p-value** |
| --- | --- | --- | --- |
| **Age, mean (SD)** | 44.7 (10.4) | 52.5 (12.0) | <0.001* |
| **Female %** | 55.5 | 23.1 | <0.001* |
| **BMI, mean (SD)** | 22.4 (3.2) | 25.1 (4.1) | <0.001* |
| Overweight % | 18.6 | 34.2 | <0.001* |
| Obesity grade I % | 1.7 | 9.0 | <0.001* |
| Obesity grade II % | 0.2 | 1.5 | 0.057 |
| Obesity grade III % | 0.0 | 0.6 | 0.020* |
| **Tobacco smoking pack-years, mean (SD)** | 8.3 (8.8) | 11.1 (14.4) | 0.146 |
| **Alcohol consumption** |  |  |  |
| Daily % | 5.2 | 10.6 | <0.001* |
| Several times/week % | 64.9 | 58.4 | 0.020* |
| Monthly/yearly % | 19.0 | 19.7 | 0.739 |
| Abstinent % | 11.0 | 11.3 | 0.851 |
| **Diastolic BP, mean (SD)** | 71.6 (5.4) | 86.1 (8.3) | <0.001* |
| **Systolic BP, mean (SD)** | 110.1 (6.7) | 131.3 (11.5) | <0.001* |
| **Pulse pressure, mean (SD)** | 38.5 (5.6) | 45.2 (8.7) | <0.001* |
| **Self-reported poor health, %** | 14.4 | 18.6 | 0.041* |
| **Other CVD (excluding hypertension) %** | 14.6 | 26.4 | <0.001* |
| **Measured renal deficiency (%)** | 0.64 | 1.73 | 0.141 |
| **Diabetes %** | 0.9 | 2.8 | 0.031* |
| **Sleep apnea %** | 0.9 | 8.8 | <0.001* |

*p < 0.05 = statistically significant (*)*
